# Development of a quantitative antigen assay to detect coccidioidal chitinase-1 (CTS1) in human serum

**DOI:** 10.1101/2021.05.14.21257226

**Authors:** Francisca J. Grill, Thomas E. Grys, Marie F. Grill, Alexa Roeder, Janis E. Blair, Douglas F. Lake

## Abstract

**Background:** Coccidioidomycosis is often diagnosed with a collection of tests that rely on the patient’s ability to mount an immune response to the fungus (antibody-based diagnostics), making diagnosis of this infection challenging. Here we present an antigen-based assay that detects and quantifies coccidioidal chitinase-1 (CTS1) in human serum.

**Methods:** An inhibition-based enzyme-linked immunoassay (ELISA) was developed that utilizes a monoclonal antibody specific for coccidioidal CTS1. CTS1 was quantified in commercial antigen preparations using recombinant CTS1 as a standard. Sera from 192 individuals from an endemic area were tested which included 78 patients (40.6%) with proven or probable coccidioidomycosis.

**Results:** The quantity of CTS1 in diagnostic commercial antigen preparations from different suppliers varied. CTS1 antigenemia was detected in 87.2% of patients with proven or probable coccidioidomycosis. Specificity was determined to be 96.94% using serum from individuals who reside in the Phoenix, Arizona area who did not have coccidioidomycosis. Levels of CTS1 correlated with low- and high-titer serology from patients with a coccidioidomycosis diagnosis.

**Conclusions:** Since the CTS1 inhibition ELISA described in this report does not depend on the host immune response, it is a promising diagnostic tool to aid in diagnosis and disease monitoring of coccidioidomycosis.

**Summary:** Diagnosis of coccidioidomycosis often relies on the host’s ability to mount an immune response. Here we present an antigen-based assay that detects and quantifies coccidioidal chitinase-1 in human serum to diagnose coccidioidomycosis independent of host immune status.

## Introduction

Coccidioidomycosis, or Valley Fever (VF), is an infection caused by the pathogenic fungi *Coccidioides immitis* and *Coccidioides posadasii* which are nearly identical in their morphology and physiology[1]. *Coccidioides* spp. are present in the southwestern and western United States, northern Mexico, and small areas in Central and South America[2–4]. In 2018, over 15,000 cases of VF were reported in the United States with a majority in Arizona and California, both of which have experienced a significant increase in incidence since 2014[5–7]. Infection may be asymptomatic or manifest as a pneumonia difficult to distinguish from community-acquired viral or bacterial pneumonia which may lead to inappropriate treatment[8,9]. In a small but significant number of cases, extra-pulmonary dissemination of the fungus occurs resulting in significant morbidity, need for long-term antifungal therapy, and sometimes in death[10,11].

*Coccidioides* spp. are dimorphic fungi that exist in two forms dependent on their environment, growing as a spherule (yeast) form in vertebrate hosts and septate mycelia in the desert soil. Under dry conditions in the environment, spore-like arthroconidia form, become easily aerosolized, and are inhaled into the lungs of a human or animal host, where the fungus transforms into the spherule growth form. As spherules grow in the host, endospores form inside, increasing the size of the spherules. Spherules then rupture and release hundreds of endospores, each capable of developing into a new spherule that undergoes the same process[12]. Chitin has been shown to be a major component in the cell wall of spherules[13] and its presence increases significantly during the beginning of the endosporulation process, likely as a result of an upregulation of chitin synthases[14]. This surge in chitin production is then diminished with the progression of endospore formation and release, at which time chitinases are detectable[10,14]. Two chitinase (CTS) genes, *cts1* and *cts2*, have been characterized and the product of the former, CTS1, is a 48-kDa protein that plays a role in weakening the spherule cell wall prior to endospore release[15–17]. In culture, the presence of CTS1 is shown to decrease shortly after endosporulation due to an upregulation of proteases[10,18], so the process of spherule growth and endosporulation can restart. CTS1 is also known as the “CF” antigen used in serodiagnostics, namely complement fixation (CF) assays and its immunodiffusion correspondent (IDCF)[16,17]. It has since been produced recombinantly by several groups, and truncations of CTS1 have been explored to map antibody-binding sites to increase sensitivity and specificity for detecting antibodies to *Coccidioides* in patients[19–23].

Even though the CTS1/CF antigen system is commonly used in antibody-based diagnostics it can take weeks to months after onset of symptoms to detect an immune response, which may be further delayed or even absent in immunocompromised patients[24]. Currently, a constellation of diagnostic assays and clinical findings are utilized to diagnose VF. Serologic tests that detect patient antibodies to *Coccidioides* spp. antigens are the most widely used and include enzyme immunoassays (EIA), immunodiffusion (ID) and complement fixation (CF)[25]. EIA and ID utilize reagents that can distinguish between IgM and IgG antibodies, while CF antigen is used to quantify the antibody response and measure disease progression and/or resolution[9,26]. The sensitivity and specificity of these assays in diagnosis have been evaluated by multiple groups[24,27–32], and although helpful, the sensitivity of serology is still lacking, especially in early infection and in immunosuppressed individuals[24]. Specificity can also be a problem. As disease resolves, the antibodies detected by clinical assays wane. Alternatively there may be complete resolution of the fungus, with a long period of detectable antibody, or incomplete resolution (latent infection), with continued detection of antibodies. Unlike many serologic assays for infectious diseases, IgG is not a marker of distant infection, but could represent recent or latent infection[31]. Thus, while serology continues to be a mainstay diagnostic tool, it is an incomplete and imperfect approach. Morphological and growth-based diagnostics such as microscopy and culture, respectively, exist. However, these methods are also lacking in sensitivity and the latter poses a risk to laboratory personnel despite being considered the gold-standard of diagnosis[9,33]. Polymerase chain reaction (PCR) has also been explored, however it demonstrates a sensitivity similar to that of culture[34,35]. This experience has therefore highlighted the need for a sensitive antigen-based diagnostic test that detects and/or measures direct biomarkers from *Coccidioides* spp. as opposed to a patient response. The use of EIA for detection of coccidioidal antigens has been approached previously using spherule lysate[36], and more recently using galactomannan[37]. No follow up evaluations of the former have been explored, but the latter has shown value in diagnosing more severe-forms of disease such as coccidioidal meningitis[37,38]. A recent publication affirmed the modest incremental value of performing the *Coccidioides* galactomannan antigen assay, though the assay was positive for only a minority of non-disseminated cases[32].

In this report, we describe a new enzyme-linked immunoassay (ELISA) that measures coccidioidal CTS1. Evaluation of this assay included measuring levels of CTS1 in commercial antigen preparations and testing 192 serum samples from individuals residing in an endemic area (Phoenix, AZ). The inhibition ELISA demonstrated 96.94% specificity and 87.18% sensitivity when compared to an algorithm that includes histology, culture, clinical symptoms, radiographic and serologic findings. However, since the assay is not dependent on the host immune response, the ability to detect the presence of fungal antigen in serum could provide a definitive diagnosis for patients suspected of having coccidioidomycosis.

## Methods

### Production and purification of recombinant CTS1

The CTS1 gene was provided to us as a kind gift from Dr. Mitch Magee (Arizona State University). We cloned CTS1 into pcDNA3.1 V5/HisA, verified its identity by sequence analysis (supplemental Figure 1) and transfected it into 293F cells (Thermo Freestyle Expression System). Seven-day supernatants were harvested and recombinant CTS1 (rCTS1) was purified on a nickel column via a C-terminal histidine tag. Purified protein was quantified using a BCA protein assay kit (Thermo Scientific) and frozen at −80°C in 250ul aliquots at 1 mg/ml.

**Figure 1:**
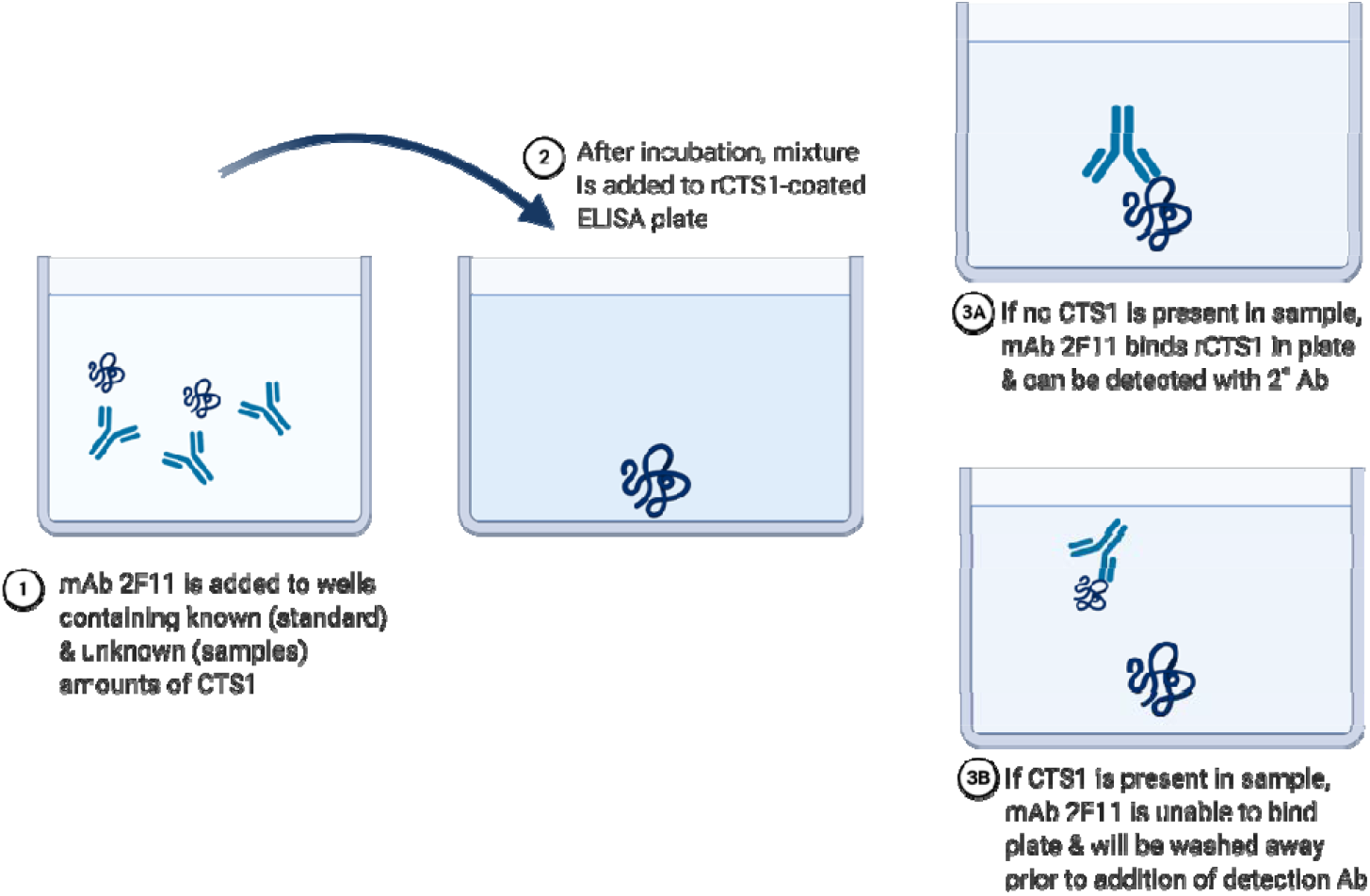
Diagram of inhibition assay used for quantification of CTS1. Biontinylated monoclonal antibody 2F11 is incubated with patient serum at different dilutions for one hour. After incubation, mixture is added to an ELISA plate that has been coated with rCTS1 and allowed to incubate. If CTS1 is present in the sample being tested, it will block the antibody from being able to bind rCTS1 in the ELISA plate, generating low signal. If CTS1 is not present, the antibody will bind rCTS1 in the ELISA plate, generating high signal. Schematic was created with BioRender.com.

### Monoclonal antibody generation and purification

Mice were immunized and boosted with rCTS1 mixed with Magic Mouse Adjuvant (Creative Diagnostics) under an IACUC approved protocol at Mayo Clinic. Anti-CTS1 antibody titers were monitored by rCTS-1-coated 96-well plates. When a sufficient antibody titer was reached (>1:32,000), mice were sacrificed and spleens were processed into single cell suspensions. Splenocytes were fused with myeloma cells (P36X3Ag8.653) by a standard hybridoma generation technique[39]. Briefly, splenocytes were fused with P3X63Ag8.653 myeloma cells at a ratio of 1 splenocyte: 2 myeloma cells using 50% PEG (Sigma-Aldrich). The fused cells were resuspended in hypoxanthine-aminopterin-thymidine (HAT) selective medium (Sigma-Aldrich) and plated at 50,000 splenocytes per well in a 96-well plate. Plates were incubated at 37°C with 5% CO2 for ten days. Supernatant was collected and tested by indirect ELISA for antibodies against rCTS1. Positive wells were subcloned by limiting dilution of one cell per well and re-screened using the same procedure after ten days. Positive subclones were cultured in 10% FBS cDMEM for antibody purification by protein A/G (Thermo Scientific) chromatography. Multiple mAbs against rCTS1 were identified, but one in particular, 2F11, performed well in our inhibition assay.

### Western blot

rCTS1 was either treated with PNGaseF or untreated and subjected to SDS-PAGE under reducing conditions using 12% polyacrylamide gels at 140 V for 60 minutes (Bio-Rad) followed by staining with Coomassie Blue dye. For Western blot analysis, electrophoresed proteins were transferred to a PVDF membrane using a western blot apparatus (Bio-Rad). After transfer, membranes were blocked in 1% BSA for at least one hour followed by incubation with anti-CTS1 mouse monoclonal antibody 2F11 at 2 ug/ml in 1% BSA in PBS for 1 hour. Membranes were then washed three times with tris-buffered saline, 0.1% Tween-20 (TBST) followed by addition of peroxidase-conjugated goat anti-mouse IgG antibody (Jackson ImmunoResearch) at a dilution of 1:5000 for 45 minutes. Membranes were subsequently washed four times with TBST followed by addition of NBT/BCIP (Thermo Scientific) substrate for development.

### Inhibition ELISA

The first step of the inhibition assay requires pre-incubating a biofluid potentially containing *Coccidioides*-produced CTS1 with a calibrated concentration of 2F11 anti-CTS1 mAb. Then, the solution is transferred to rCTS1-coated ELISA plates (Figure 1). A sample that does not contain any CTS1 would result in 2F11 mAb binding to rCTS1 coated on the plate, whereas a sample containing CTS1 would have already bound some or all of the 2F11 mAb, inhibiting 2F11 mAb from binding to plate-bound rCTS1. A standard of rCTS1 was run with each test so that CTS1 in the biofluid could be compared at an appropriate dilution along the linear portion of the standard curve.

Commercial antigen preparations and human sera were tested undiluted and at 2-fold dilutions in 1% BSA. The assay was performed using a 96-well flat bottom plate (Corning) coated with rCTS1 at 2ug/ml for 75 minutes at 37°C followed by blocking overnight in 1% BSA in 1X PBS at 4°C. The following day, rCTS1 standard was spiked in a 96-well U-bottom plate (Corning) at a known concentration (16 ug/ml) into 1% BSA or normal donor serum followed by ten 2-fold dilutions into 1% BSA. Biotinylated 2F11 mAb was added to either commercial antigen preparations or sera at an equal volume such that the final concentration of 2F11 was 0.1 ug/ml, final dilution of standard 8ug/ml, and final dilution of samples 2-fold greater than starting dilution (e.g., starting dilution of 1:2 became 1:4). Samples were mixed by gently tapping the 96-well U-bottom plates followed by incubation at room temperature for one hour. Samples were then transferred to rCTS1-coated ELISA plates and incubated an additional hour. ELISA plates were then washed three times with 1X PBS, 0.05% Tween-20 (PBST), followed by addition of Streptavidin-HRP (BD Biosciences) at a dilution of 1:5000 and incubated for 45 minutes. The plates were washed three times in PBST, then developed with TMB substrate (BD Biosciences) for ten minutes. Sulfuric acid 0.16M was added to stop development and the plate was read at 450nm.

### Assay limits and precision

A standard curve for quantification of CTS1 was generated by spiking a known concentration of rCTS1 into either 1% BSA or normal donor serum followed by two-fold dilutions into 1% BSA. The limit of blank (LOB) and limit of detection (LOD) were determined by measuring the standard curve in triplicate. The LOB was calculated by taking the mean OD value of triplicate blank samples and subtracting 1.645 times their standard deviation[40]. Subtraction instead of addition of standard deviation was used due to the reverse nature of our standard curve (low optical density corresponds to high concentration of CTS1 while high optical density corresponds to low concentration of CTS1). The LOD was determined by using the LOB and replicates of the CTS1 standard with concentrations that approached the LOB with the following equation: LOD = LOB – 1.645(SD_low concentration sample_)[40]. Once again, subtraction of SD was utilized in place of addition due to the reverse nature of our standard curve. The LOD optical density value was then back calculated to a concentration of CTS1 to account for minor day-to-day variation. This process was repeated across three days for both rCTS1 spiked into 1% BSA and normal donor serum, with dilutions into 1% BSA. The average LOD concentration is reported. The back-calculated concentrations from these replicates were used to calculate intra- and inter-assay precision of the CTS1 standard (supplemental Table 1).

### Clinical specimens

Human sera were obtained under ASU IRB 0601000548 and Mayo Clinic IRB 12-000965. All samples with at least one *Coccidioides*-related diagnostic test were collected between May-September 2018 and stored <-20°C until use. Samples were tested in dilution replicates of 1:2 - 1:32. If dilution replicate did not allow for quantification within the assay limits, samples were re-tested at higher dilutions. The status of each patient at time of sampling was categorized as “Proven”, “Probable”, “Possible”, or “not coccidioidomycosis” using European Organization for Research and Treatment of Cancer (EORTC) criteria for endemic mycoses, with clarifying criteria for Probable and Possible, based on our published clinical composite reference standard[31,41]. Briefly, patients who had *Coccidioides* identified by culture, histopathology, or PCR were classified as Proven. Patients with concurrent clinical findings (including either radiology findings[31] or symptoms[31]) along with positive serology (antibodies) for *Coccidioides* were classified as Probable. Our group classified anyone with either relevant clinical findings or positive serology, but not both, as possible coccidioidomycosis, however there is not a clear consensus about what criteria must be met for this classification[41]. Since the true nature of the Possible category cannot be known, these patients were excluded from the sensitivity and specificity analysis. Any patient who did not have Proven, Probable, or Possible coccidioidomycosis, or was diagnosed with a different illness, was classified as Not Coccidioidomycosis.

### Statistical analysis

Receiver operating characteristic analysis was used to determine the cutoff for positivity as well as estimate sensitivity and specificity. Samples with OD values below the LOD were assigned a value of 0 ug/ml for statistical analysis.

## Results

Recombinant CTS1 was electrophoresed with and without pre-treatment with PNGase F (Figure 2A). rCTS1 appears as a doublet band, which is presumed to be a glycosylation since rCTS1 appears as a single band after deglycosylation with PNGase F (∼35 kDa). Although multiple mAbs were identified from anti-CTS1-secreting mouse hybridomas, one mAb in particular, 2F11, demonstrated binding in both ELISA and western blotting. A western blot of 2F11 reacting with rCTS1 is shown in Figure 2B. This antibody was used to develop the inhibition assay. Dilutions of rCTS1 were used in an empirical approach to establish the limit of blank, which was then used to calculate the limit of detection of *Coccidioides* rCTS1, 155 ng/ml (SD 0.022 µg/ml).

**Figure 2.**
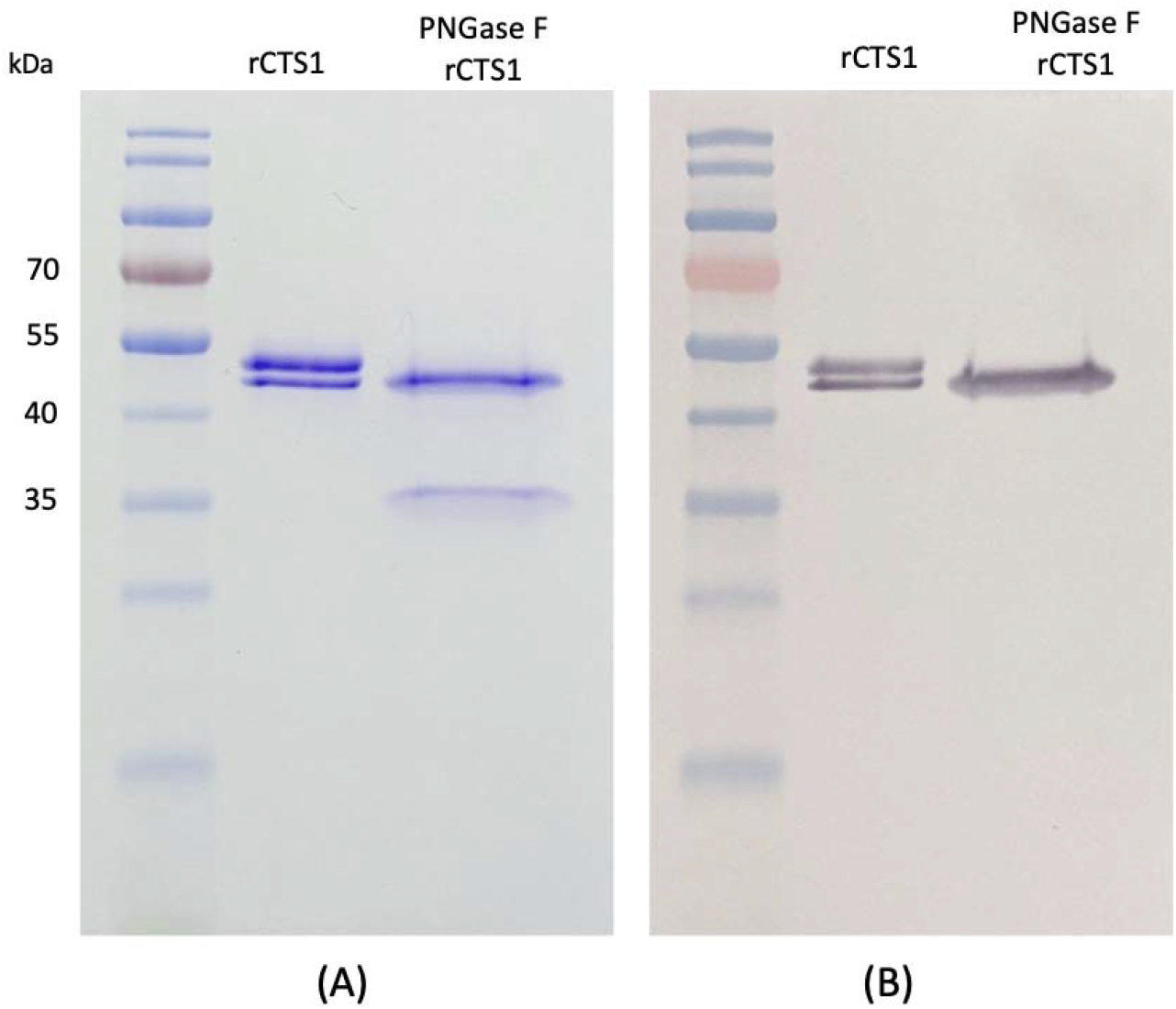
(A) Coomassie blue-stained SDS-PAGE of rCTS1 and PNGase F-treated rCTS1. (B) Western blot probed with 2F11 mAb.

The first step of characterizing the assay was to test it against commercially available antigen preparations. These included *Coccidioides* CF Antigen Concentrate (IMMY, Norman, OK), *Coccidioides* IDCF Antigen (IMMY, Norman, OK), *Coccidioides* “F” Antigen for Immunodiffusion (Meridian Biosciences, Cincinnati, OH), *Coccidioides immitis* Antigen F (Gibson Bioscience, Lexington, KY). The quantity of CTS1 in each was 2.79 ug/ml, 4.04 ug/ml, 5.29 ug/ml, and 5.88 ug/ml, respectively (Figure 3). Commercially available antigen preparations for *Aspergillus, Blastomyces*, and *Histoplasma* ID assays (IMMY, Norman, OK) were also tested, but no CTS1 was detected in non-coccidioidal antigen preparations, demonstrating the specificity of 2F11 mAb (Figure 3).

**Figure 3.**
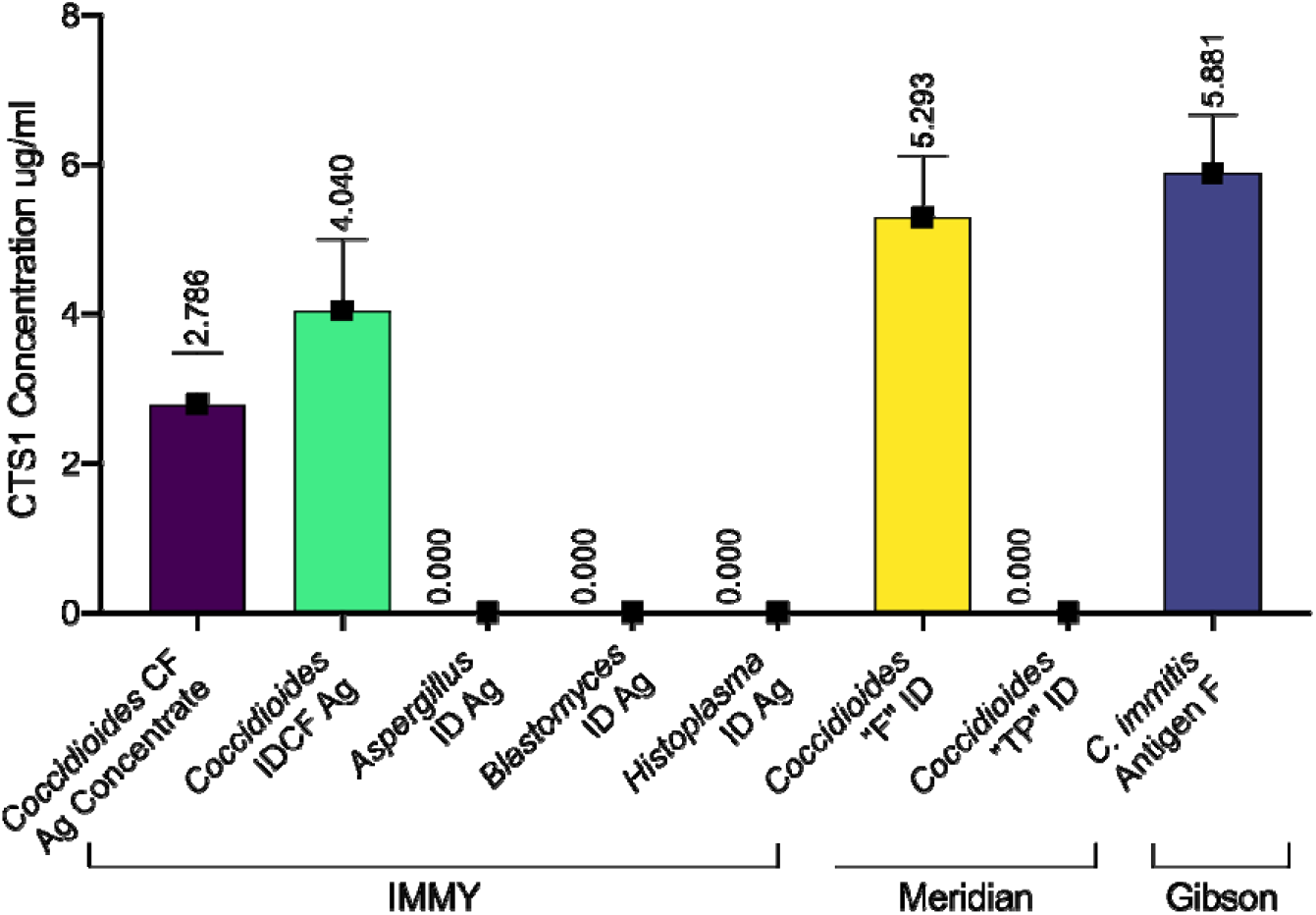
Quantification of CTS1 in different commercially available antigen preparations used in fungal diagnostics. Brackets at the bottom designate manufacturers of each antigen preparation. For values below 0.155 ug/ml (the analytical limit of detection), the assigned value was zero.

These initial findings led us to investigate if CTS1 was detectable in patient serum using the same inhibition ELISA format. We tested 192 pre-existing serum samples from patients who had at least one coccidioidomycosis-related diagnostic test performed. Patients were classified as Proven, Probable, Possible, or Not Coccidioidomycosis, based on EORTC criteria[41] and refined based on our previously described clinical schema[31]. Seventy-eight (40.6%) patients had Proven or Probable coccidioidomycosis, 16 (8.3%) had Possible coccidioidomycosis, and the remaining 98 (51.0%) did not have coccidioidomycosis (Figure 4A). Of the 98 samples classified as Not Coccidioidomycosis, 9 (4.7%) were diagnosed with a different fungal infection (*Aspergillus* n=6, *Candida* n=3), further demonstrating specificity of the assay against these filamentous and yeast form fungi. The mean concentration of CTS1 in dilution replicates for all samples tested is shown in Figure 4B and 4C. Of the 78 patients with Proven or Probable coccidioidomycosis, 51 had a positive complement fixation titer ranging from 1:2 to 1:256. The quantity of CTS1 detected in these patients in relation to CF titer is shown in Figure 4D.

**Figure 4.**
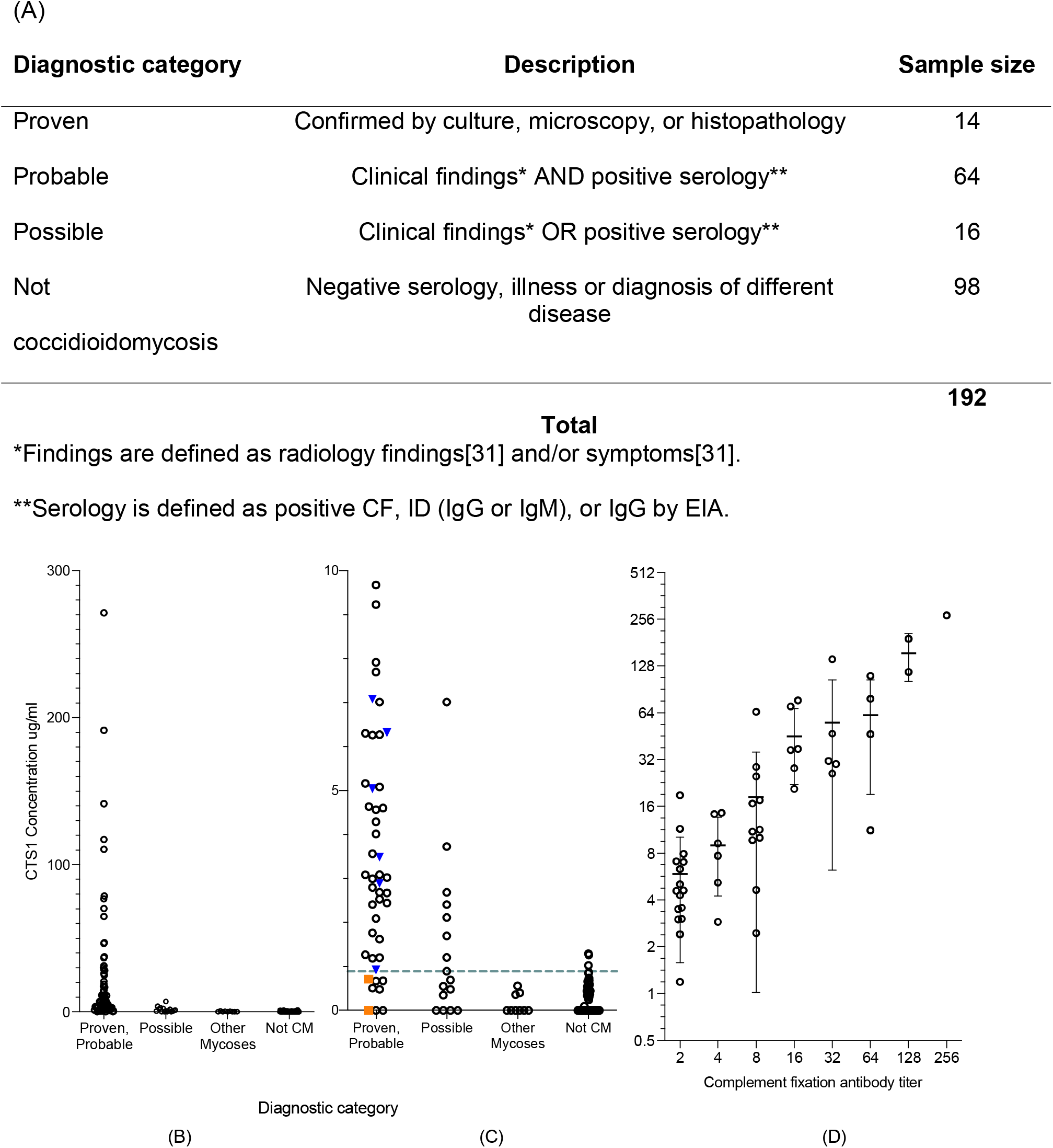
(A) Composition of serum samples tested and sorted by diagnosis category. (B) Concentration of CTS1 detected in all patient serum tested, separated by diagnostic category. Because a high concentration of CTS1 was detected in some patients, (C) displays data using a scale of 0-10 ug/ml for better visualization of distribution of concentrations. Blue inverted triangles represent patients with Proven diagnosis, while orange squares indicate patients with Proven coccidioidomycosis whose disease appeared to be resolved at time of specimen collection (no symptoms and negative serology, see supplemental Table 2). A dotted line represents the cutoff value for positivity at 0.90 ug/ml. (D) Correlation of CTS-1 levels with serologic antibody titers determined by complement fixation. Individual dots show quantity of CTS1 determined by ELISA in 51 patients with a positive complement fixation titer (grouped by CF titer on x-axis). Average quantity by titer is represented with a line, with error bars illustrating standard deviation (with the exception of CF titer 256 group, which only included 1 patient).

A receiver operating characteristic curve (ROC) was plotted using patients with Positive or Probable coccidioidomycosis as true positives and Not Coccidioidomycosis patients as true negatives (Figure 5) while excluding the Possible group, as their status is unknown. The area under the receiver operating characteristic curve was 0.9428 (SE, 0.02137; 95%CI, 0.9009-0.9847; P<0.0001) using a positive cutoff value of 0.90 ug/ml, resulting in a sensitivity and specificity of 87.18% and 96.94%, respectively.

**Figure 5.**
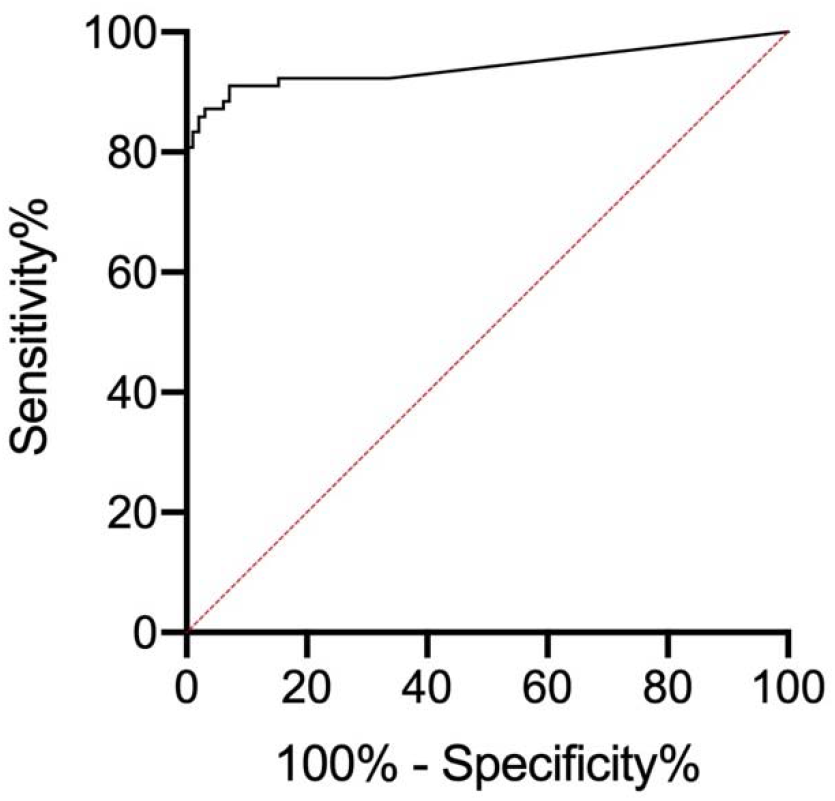
Receiver operating characteristic (ROC) plot for CTS1 quantification for the diagnosis of coccidioidomycosis. Patients with proven and probable coccidioidomycosis represent the true positive group while patients categorized as not coccidioidomycosis make up the true negative group. Patients classified as possible coccidioidomycosis were excluded from this analysis. The area under the curve (AUC) was 0.9428 (SE, 0.02137; 95% CI, 0.9009-0.9847; P<0.0001). With a cutoff concentration of 0.90 ug/ml, the sensitivity is 87.18% and the specificity is 96.94%.

## Discussion

Current diagnostic methods for coccidioidomycosis rely on a constellation of clinical, radiologic and laboratory findings that are often conflicting and inconsistent. However, their collective use is the current accepted standard used to inform clinical decisions, and often repeated testing over time is needed to clarify patient status. Further complicating diagnosis is that early treatment of coccidioidomycosis can abrogate serocoversion[42], which can make definitive diagnosis nearly impossible in some patients with other co-morbidities that have overlapping symptoms. Various CTS1 antigen preparations have been used in antibody-based diagnostic assays for over six decades. Although CTS1 may be an accepted serologic target, antibody responses in individuals infected with *Coccidioides* spp. are often delayed or even absent, especially in immunocompromised patients[24]. However, if a fungal antigen could be detected in a biofluid, a definitive diagnosis could be made, independent of the host immune response.

To begin construction of the antigen detection assay, we first generated mouse mAbs to rCTS1. During screening, mAb 2F11 was particularly notable because it bound both soluble and plate-bound rCTS1, making it an ideal antibody for an inhibition ELISA. Conditions were established and optimized for the inhibition assay diagrammed in Figure 1 in which a biofluid (serum or commercial antigen preparation) potentially containing coccidioidal CTS1 was pre-incubated with biotinylated mAb 2F11. The quantity of CTS1 in the biofluid proportionally inhibited the binding of mAb 2F11 to the rCTS1-coated plate as quantified using a standard curve generated with known concentrations of rCTS1.

Complexity and inconsistency in the current diagnostic testing regimen make evaluation of a new diagnostic tool challenging. Several groups have proposed criteria to classify patients with varying certainties of coccidioidomycosis infection to aid with this challenge. We opted to categorize the patients included in this study using the general definitions offered by the EORTC and the Mycoses Study Group[41] who provide criteria for proven and probable coccidioidomycosis, but acknowledge disagreement about the parameters of “possible” disease, Therefore, we included additional detail to define probable and possible based on our previous work[31]. Lastly, we excluded patients categorized as possible from our sensitivity and specificity analysis, since their true status is unknown. The results show that the use of our antigen EIA provides sensitivity and specificity comparable only to the use of multiple existing serodiagnostic tests[24,31,32] with the added advantage of detecting elements of the fungus itself rather than a host response to infection. The test we describe could allow for earlier diagnosis, which is crucial for proper management of patients. Some patients take many weeks to become antibody-positive[24] and are given one or more courses of unnecessary antibiotics, posing risk for patients and contributing to antimicrobial resistance.

Another use of this assay is to measure the CTS1 present in antigen preparations, and potential lot to lot variability. While the relative amounts of CTS1 between commercial products is somewhat irrelevant, it is essential to have consistent reagents for diagnostic methods. Importantly, *Histoplasma* and *Blastomyces* antigen ID preparations from IMMY did not cross-react on our assay, demonstrating a critical feature of analytical specificity of the assay. Nine patients had other fungal infections and were negative by the assay, suggesting that the assay has good clinical specificity. More samples from patients with other fungal infections will need to be tested to further characterize clinical specificity of the assay.

We must also acknowledge that the samples tested in this study are a single point in time from individuals at different stages of disease with a wide range in time since diagnosis, time since treatment initiation, and/or time since symptom resolution. In some cases, the course of disease led to evidence that would categorize the patient as a proven case. We categorized patient samples with information available at the time of blood draw. Additional context from future events is available in Supplemental Table 2 for thirty patients. In one example, Patient 3 was characterized as Probable due to innumerable pulmonary nodules and positive serology by CF (1:2), ID, and EIA. The patient’s serum contained 7.086 ug/mL of CTS1 antigen at that time. The patient was initiated on antifungal treatment 4-5 weeks later, however 11 months later returned a positive PCR, confirming proven infection. It would be difficult to know whether this patient was sufficiently treated, given this presentation. With our assay, trending of antigenemia may correlate with clinical response and/or response to treatment. To investigate this possibility, we plan to test samples longitudinally with close correlation to clinical symptoms and other diagnostic tests.

The one other antigen assay described for *Coccidioides* was recently shown to be positive in only 38.6% of sera from immunocompetent patients, and only 37.1% of pulmonary cases[32]. That assay is also positive more often in disseminated cases, which typically have other indicators of infection already. The assay reported here has the potential for similar or better performance versus any single antibody assay or combination of antibody assays, and has the distinct advantage of directly measuring concentrations of fungal antigen in a blood sample. Although additional prospective studies are needed to further evaluate and define the role of this test in the clinical laboratory, it may hold promise as a useful diagnostic tool for a disease that can be challenging to diagnose.

## Data Availability

Data is provided in manuscript (no external links).

## Acknowledgements

We would like to thank the Mayo Clinic Department of Laboratory Medicine and Pathology for their support in acquiring samples to use in this study.

## Author contributions

Francisca J. Grill: Investigation, Methodology, Data Collection, Retrospective Chart Review, Writing – Original Draft and Editing

Thomas E. Grys: Conceptualization, Methodology – Sample Classification, Resources, Writing – Reviewing and Editing

Marie F. Grill: Methodology – Sample Classification, Writing – Reviewing and Editing

Alexa J. Roeder: Sample Collection, Retrospective Chart Review

Janis E. Blair: Methodology – Sample Classification, Writing – Reviewing and Editing

Douglas F. Lake: Conceptualization, Methodology, Resources, Supervision, Writing – Reviewing and Editing

