## Supplemental figures for "Development of a quantitative antigen assay to detect coccidioidal chitinase-1 (CTS1) in human serum"

Supplemental Figure 1. Sequence confirmation of recombinant CTS1 by mass spectrometry. Tryptic digest of recombinant CTS1 was analyzed by LC-MS/MS. Spectra were searched against a combined FASTA database of *Coccidioides* spp. proteomes (obtained from Uniprot) using Sequest in Proteome Discoverer v1.4.1.14 (Thermo). Endochitinase-1 (accession no. P0CB51) was identified with 82.44% coverage as seen in highlighted regions. The highlight color corresponds to peptide confidence, with green indicating high-confidence peptides and red indicating low-confidence peptides.

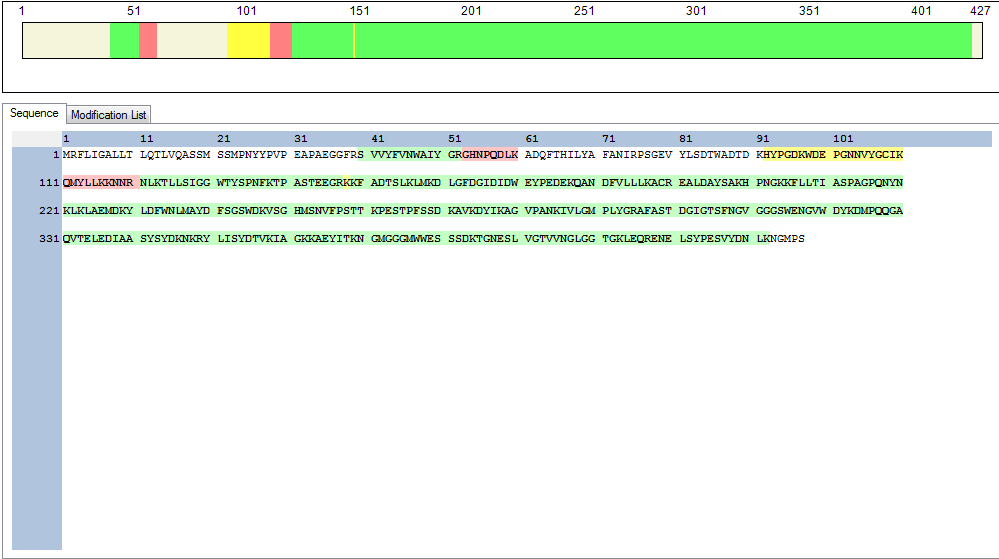

Supplemental Table 1. Intra- and inter-assay variation of the Coccidioidal CTS1 quantification ELISA. Standard curves were run in triplicate on six plates over multiple days. Intra-assay coefficient of variation (CV) was calculated for back-calculated concentrations within each plate and the average intra-assay CV across all plates for each standard is the value shown. Inter-assay CV was calculated using the mean back-calculated concentration for each standard across all plates (mean of triplicate means). CV% is calculated by dividing the standard deviation by the mean and converting to a percentage (x100).

| **Parameter** | **Standard (ug/ml)** | **CV (%)** |
| --- | --- | --- |
| Intra-assay | 8.000 | 10.31 |
|  | 4.000 | 6.37 |
|  | 2.000 | 7.42 |
|  | 1.000 | 8.66 |
|  | 0.500 | 6.91 |
|  | 0.250 | 10.65 |
| Inter-assay | 8.000 | 10.39 |
|  | 4.000 | 8.85 |
|  | 2.000 | 9.91 |
|  | 1.000 | 9.59 |
|  | 0.500 | 8.04 |
|  | 0.250 | 9.21 |

Supplemental Table 2. Additional information on 30 samples. Samples in this table include those with proven and probable coccidioidomycosis (CM) who were below the cutoff for positivity or were categorized as probable but later would have been proven; all samples in the possible CM category; and patients classified as not CM who had detectable antigen.

| Patient | History, presentation |  | Radiology^#^ | Serological test | | | Antigen concentration, ug/ml | Subsequent disease course | Likely best-fit classification |
| --- | --- | --- | --- | --- | --- | --- | --- | --- | --- |
|  |  | Category |  | **CF** | **ID** | **EIA** |  |  |  |
| 1 | Dx with proven DCM 30 months prior to specimen collection. 24 months of antifungal tx. No symptoms reported at time of specimen collection | Proven | 2cm nodule | Neg | IgG - IgM - | IgG - IgM - | 0.709 | 15 months later IgG + by ID | Resolving/  Controlled |
| 2 | Dx with proven CM 12 months prior to specimen collection. Subsequent antifungal tx. No symptoms reported at time of specimen collection | Proven | Nodule with stable surrounding micronodules | Neg | IgG - IgM - | IgG in. IgM - | <0.155 |  | Resolving/  Controlled |
| 3 | Cough, dyspnea at time of specimen collection | Probable | Innumerable small pulmonary nodules | 2 | IgG + IgM + | IgG + IgM - | 7.086 | 11 months later culture and PCR positive for *Coccidioides* | Chronic |
| 4 | Dx with CM 3 months prior to specimen collection. Initiated antifungal tx. Fatigue, chest pain, dyspnea with exertion at time of specimen collection | Probable | Small nodule | Neg | IgG - IgM + | IgG + IgM + | 0.505 | 2 months later chest x-ray is clear, IgM + on ID. 4 months later all serology negative | Resolving/  Controlled |
| 5 | Feels ill, feverish, general malaise | Probable | 1 month after specimen collection shows nodule with ground glass opacities | Neg | IgG - IgM + | IgG + IgM - | <0.155 | Blood culture positive for Candida. 4 months later negative by ID | Resolving/  Controlled |
| 6 | Severe fatigue, shortness of breath. Symptoms have been present for 6 months | Probable | Lungs appear clear. X-ray 6 months prior shows two nodules. | Neg | IgG - IgM - | IgG + IgM - | <0.155 | Remained IgG + by EIA 5 months after specimen collection | Resolving/  Controlled |
| 7 | Dx with CAP suggestive of CM 9 months prior to specimen collection. Deep cough with yellow-green sputum and shortness of breath at time of specimen collection. Family came down with similar acute respiratory illness after international travel but have recovered. | Probable | 1cm nodular density | Neg | IgG - IgM - | IgG + IgM + | <0.155 | No future serology performed | Resolving/  Controlled |
| 8 | Kidney transplant evaluation 9 months prior to specimen collection, positive for CM IgM by ID and IgG by EIA. No hx of symptomatic CM. Asymptomatic at time of specimen collection | Probable | Calcified granuloma, numerous tiny nodules appear partially calcified. | Neg | IgG - IgM + | IgG + IgM - | <0.155 | Subsequent serology remains the same | Resolving/  Controlled |
| 9 | Fatigue, dizziness, tension in head. Hx of Sjogren’s syndrome and RA | Probable | Vague nodular density, could represent pulmonary nodule, no comparison available | Neg | IgG + IgM - | IgG + IgM - | 0.6682 | Remained IgG + by ID 3 months later, then negative 7 months later | Undetermined |
| 10 | Hx of AML. Hospitalized 1 month prior to specimen collection due to fatigue and weakness. Antifungal tx initiated due to positive IgG by EIA. Reports fatigue, weakness and mild cough at time of specimen collection | Probable | Multifocal ground-glass opacities and consolidative nodularities significantly improved | Neg | IgG - IgM + | IgG + IgM - | <0.155 | Remained IgM + by ID one month later, then negative 3 months later | Resolving/  Controlled |
| 11 | Went to urgent care 2 weeks prior to specimen collection, initiated on 10-day course of Levaquin, then Cefuroxime due to failure to resolve symptoms. At time of specimen collection is having fevers, night sweats, sinus congestion, cough, and chest tightness | Probable | Focal consolidation consistent with left lower lobe pneumonia | Neg | IgG - IgM + | IgG + IgM + | 0.673 | IgG + by ID 1 month later. Negative by ID but IgG + by EIA 3 months later. CF titer positive at 1:2 6 months later with subsequent increase to 1:4 | Potentially Acute infection |
| 12 | Hx of CM by positive serology 13 months prior to specimen collection, antifungal tx initiated. At time of specimen collection patient undergoing liver transplant, antifungal tx resumed. | Possible | Mild bibasilar subsegmental atelectasis | Neg | IgG - IgM eq. | IgG + IgM - | <0.155 | Subsequent serology since transplant has been consistently negative | Resolving/  Controlled |
| 13 | Symptoms 1 month prior included profound fatigue, severe headaches, palpitations, chest pain, malaise, night sweats. At time of specimen collection patient reports improvement, has completed 2 weeks of antifungal tx. | Possible | Nodular mass-like consolidation with surrounding ground-glass and numerous satellite nodules | Neg | IgG - IgM eq. | IgG - IgM - | 0.6915 | IgG + by EIA only 5 months later. Serology since then has been negative. | Resolving/  Controlled |
| 14 | Presented 3-4 months prior to specimen collection for cough, extreme fatigue, sore throat, shortness of breath. Initiated antifungal tx and noted slow improvement. Stopped antifungal tx two weeks prior to specimen collection, noticed worsening of chest discomfort and fatigue. | Possible | No abnormalities noted | Neg | IgG - IgM - | IgG - IgM in. | <0.155 | Continued IgM in. or + result by EIA only. CF and ID consistently negative. | Undetermined |
| 15 | No clinical notes about symptoms at time of specimen collection. | Possible | 2 nodular opacities, both noncalcified | Neg | N/A | IgG - IgM - | 0.349 | No previous or subsequent serology | Undetermined |
| 16 | Dx with CM 18 months prior to specimen collection due to symptoms and serology. At time of specimen collection patient has completed 18 months of antifungal tx and is asymptomatic | Possible | Chest x-ray 6 months prior to specimen collection shows decrease in size and density of nodule, no new nodules noted | Neg | IgG - IgM - | IgG - IgM in. | 0.483 |  | Resolving/  Controlled |
| 17 | Dx with CAP 2 years prior to specimen collection, resolved. Dx. With probable CM 1 year prior to specimen collection due to fatigue, fever, cough, shortness of breath. Antifungal tx for 4-6 weeks followed by resolution of symptoms. At time of specimen collection has experienced a relapse of symptoms over past two months. | Possible | Chest CT 2 months prior to specimen collection shows improvement in tree-in-bud nodules. Chest x-ray 1 month after specimen collection shows peribronchial vascular nodules. | Neg | IgG - IgM - | IgG - IgM - | 1.207 | All future serology remains negative | Resolving/  Controlled |
| 18 | Symptoms unclear/ not specified at time of specimen collection. Within 2 years prior to specimen collection has been dx with lupus erythematosus, babesia, toxoplasma, and chlamydia pneumonia | Possible | Calcified granuloma right middle lobe; nodule right lower lobe | Neg | IgG - IgM - | IgG - IgM - | <0.155 | No subsequent serology | Undetermined |
| 19 | Tx with azithromycin and prednisone 7 months prior to specimen collection for pneumonia. 6 months prior was dx with CM by positive serology but elected not to initiate antifungal tx. Specimen collected one month post sx, feels somewhat sub-fatigued | Possible | No clear abnormalities noted | Neg | IgG - IgM - | IgG + IgM - | 2.686 | No subsequent serology | Resolving/  Controlled |
| 20 | Dx with possible CM 6 months prior to specimen collection. Asymptomatic at time of collection. | Possible | Lungs appear clear | 2 | IgG + IgM - | IgG – IgM in. | 7.013 | CF titer remained at 1:2 one year after specimen collection | Resolving/  Chronic |
| 21 | Slightly increased fatigue, upper respiratory symptoms, allergies, inconsistent cough and asthma. Patient concerned about a neck lump that has increased in size over past 3 months. | Possible | 6 months prior to specimen collection no abnormalities noted. 2 weeks post specimen collection chest CT notes 2mm nodule “probably of no clinical consequence” | Neg | IgG - IgM - | IgG - IgM + | <0.155 | No subsequent serology | Undetermined |
| 22 | Dx with CM 20 months prior to specimen collection. Over 12 months of antifungal tx. Asymptomatic at time of specimen collection | Possible | Lungs appear clear | 2 | IgG + IgM - | IgG - IgM in. | 2.406 | Subsequent CF titer negative but remained IgG + by ID | Resolving/  Controlled |
| 23 | Dx with acute sinusitis one week prior to specimen collection, tx with Augmentin. At time of specimen collection sinus pressure has resolved but has lingering headache, sweats, hot flashes, sore throat. | Possible | No abnormalities noted | Neg | IgG - IgM - | IgG - IgM + | 0.893 | Subsequent serology remains IgM + by EIA | Undetermined |
| 24 | Subtle medial subclavicular swelling, asymptomatic | Possible | 19mm indeterminate nodule, not definitively identified on lateral image. No prior imaging for comparison. Lungs are otherwise clear. | Neg | N/A | IgG - IgM - | 3.729 | Subsequent serology includes EIA only, remained negative | Undetermined |
| 25 | Dx with sarcoidosis 5 months prior to specimen collection, treated with steroids but had return of symptoms with attempt to decrease steroid dose (chest pain, dry cough). | Possible | Progression parabronchial mediastinal lymph node enlargement consistent with treated or resolving sarcoidosis | Neg | IgG - IgM - | IgG in. IgM in. | 0.55 | Serology by EIA sent out one week later IgG - and IgM - | Undetermined |
| 26 | Hx of CM 5 years prior to specimen collection, also hx of COPD. At time of specimen collection has cough, occasional wheezing, oral lesion on lip. | Possible | Left perihilar lesion stable since 2017 | Neg | N/A | IgG - IgM - | 2.118 | No subsequent serology | Resolving/  Chronic |
| 27 | Patient being evaluated post-transplant | Possible | Scattered sub centimeter pulmonary nodules stable from 2016. Some atelectasis | Neg | IgG - IgM - | IgG - IgM - | 1.694 |  | Undetermined |
| 28 | Patient being evaluated for transplant | Not CM | No abnormalities noted | Neg | IgG - IgM - | IgG - IgM - | 1.292 | Subsequent serology continues to be negative. | Undetermined |
| 29 | Patient being evaluated for kidney donation | Not CM | No abnormalities noted | Neg | IgG - IgM - | IgG - IgM - | 1.295 |  | Undetermined |
| 30 | Hx of RA. Patient reports hot flashes and night sweats for past month. | Not CM | No abnormalities noted | Neg | IgG - IgM - | IgG - IgM - | 1.025 | Subsequent serology continues to be negative. | Undetermined |

\# radiology was within 3 months of specimen collection unless otherwise noted.

in = indeterminate

eq = equivocal

CM = coccidioidomycosis

DCM = disseminated coccidioidomycosis

Dx = diagnosed

Hx = history

Tx = treatment
